# Racial and Ethnic Reporting and Representation in Phase III Alzheimer’s Disease Clinical Trials in the US

**DOI:** 10.1101/2025.05.03.25326933

**Authors:** Zhuoer Lin, Ruochen Sun, Joseph S. Ross, Kien Lau, Sophia Stumpf, Xi Chen

## Abstract

**Background:** Alzheimer’s disease (AD) disproportionately affects racial and ethnic minoritized populations in the United States, yet these groups remain markedly underrepresented in clinical research. Phase III clinical trials are critical for informing regulatory decision and treatment guidelines, but the extent to which they report and include racial and ethnic diverse participants in the US context has not been systematically assessed.

**Methods:** We conducted a comprehensive retrospective review of all US-based Phase III AD clinical trials from 1997 to 2023 using the Trialtrove database, cross-referenced with PubMed, ClinicalTrials.gov, and other public sources. We analyzed long-term trends in the reporting and representation of racial and ethnic groups across the longest observation period to date.

**Results:** Of 88 identified trials, 71 (80.7%) had published data. Nearly half (49.3%) did not report any race or ethnicity information. Among those that did, most focused on White patients, with limited and inconsistent reporting for racial and ethnic minoritized groups. Median enrollment was 0.9% for Asian or Pacific Islander, 4.5% for Black (ethnicity unspecified), 7.2% for Black (non-Hispanic), 5.2% for Hispanic, and 0.4% for Native American participants, compared to nearly 90% for White participants. Only 4.2% of trials conducted subgroup analysis by race or ethnicity, and none reported detailed outcome differences. Terminology varied widely and no trials acknowledged underrepresentation or proposed corrective strategies. Notably, these patterns showed little to no improvement over time.

**Conclusions and Implications:** Racial and ethnic minoritized populations remain consistently underreported and underrepresented in Phase III AD trials in the US, limiting the generalizability of findings and risking the exacerbation of health inequities. Improving equity in AD research will require standardized reporting, inclusive recruitment practices, and intentional efforts to engage underrepresented communities.

## Introduction

Alzheimer’s disease (AD) disproportionately affects racial and ethnic minoritized populations in the United States, rendering it crucial to assess whether therapies are safe and effective across diverse groups.^1,2^ Phase III trials, the foundation for clinical guidelines and regulatory decisions, often underrepresent these populations,^3^ limiting generalizability and potentially exacerbating disparities in care. Despite well-documented differences in genetic, behavioral, and clinical factors, reporting and inclusion by race and ethnicity remains limited and inconsistent.^4^ Previous reviews often did not distinguish between trial phases, combined US and non-US trials, and focused primarily on selected bibliographic databases or trial registries, limiting their policy relevance and applicability to the US context.^4,5^ Using comprehensive trial databases, we characterized the longest trends in how race and ethnicity were reported, analyzed, and represented in US-based Phase III AD trials from 1997 to 2023, highlighting persistent gaps in reporting and representation.

## Methods

We retrospectively reviewed US-based Phase III AD clinical trials using a multi-source approach, i.e., the comprehensive Trialtrove trial database, cross-referenced with PubMed, trial registries, and other public sources for validation. Data were extracted from peer-reviewed publications, ClinicalTrials.gov, pharmaceutical reports, and conference abstracts. Trials that were incomplete or involved non-US populations were excluded (eFigure).

Primary outcomes were the reporting of race and ethnicity, terminology used, and representation of racial and ethnic minoritized populations. Secondary outcomes included subgroup analyses of treatment effects and discussion of diversity. Time trends in reporting and representation were analyzed. This study was deemed non-human subjects research by Yale University’s Ethics Committee; and followed the STROBE reporting guideline.

## Results

Of 88 US-based Phase III AD clinical trials reported from 1997-2023, 71 (80.7%) had published data available, with 52 (59.1%) appearing in peer-reviewed journals. Nearly half (49.3%) did not report patient race or ethnicity. Among those that did, most focused on White patients, while other groups were underreported. Specifically, only 15.5% of trials reported data on Asian/Pacific Islander patients, 28.2% on Black patients, 18.3% on Hispanic patients, and 2.8% on Native American patients. Only 4.2% of trials conducted any subgroup analysis by race and ethnicity; none reported detailed outcome differences by group (**Table**).

**Table 1.**
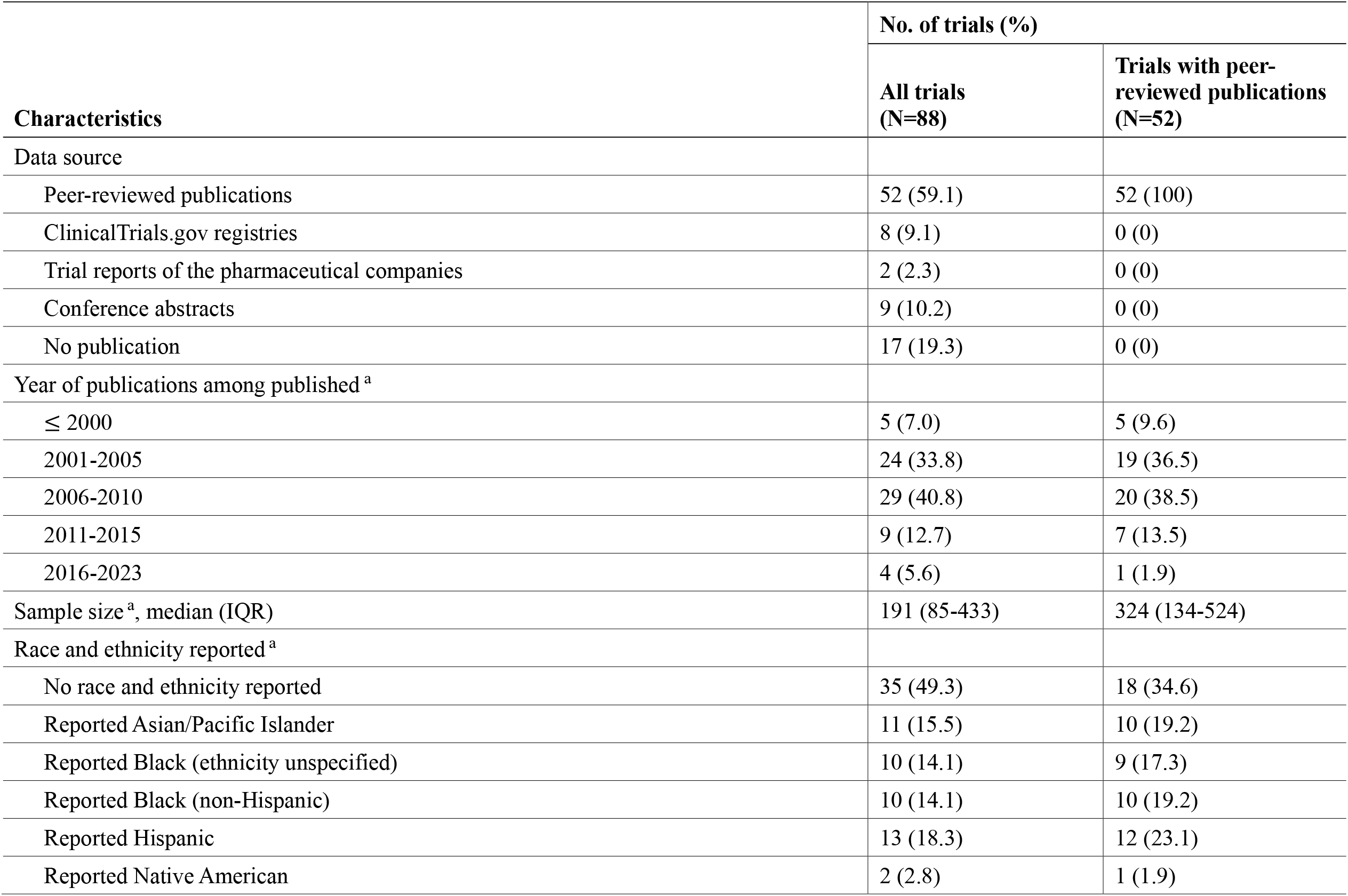
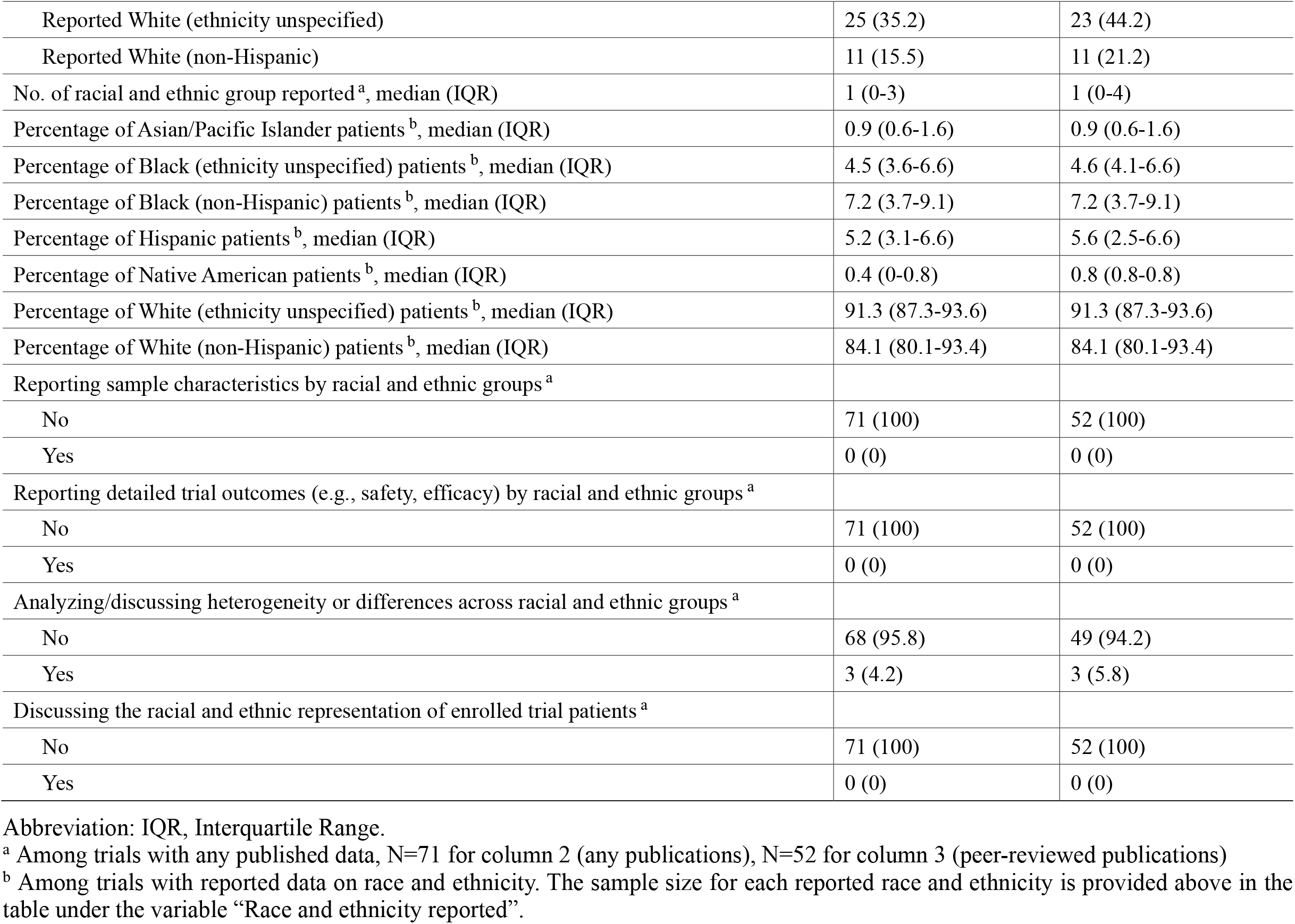
Characteristics and Racial/Ethnic Reporting and Representation in Phase III Alzheimer’s Disease Clinical Trials in the US, 1997-2023 (N=88)

| **Characteristics** | **No. of trials (%)** |  |
| --- | --- | --- |
|  | **All trials (N=88)** | **Trials with peer-reviewed publications (N=52)** |
| Data source |  |  |
| Peer-reviewed publications | 52 (59.1) | 52 (100) |
| ClinicalTrials.gov registries | 8 (9.1) | 0 (0) |
| Trial reports of the pharmaceutical companies | 2 (2.3) | 0 (0) |
| Conference abstracts | 9 (10.2) | 0 (0) |
| No publication | 17 (19.3) | 0 (0) |
| Year of publications among published <sup>a</sup> |  |  |
| ≤ 2000 | 5 (7.0) | 5 (9.6) |
| 2001-2005 | 24 (33.8) | 19 (36.5) |
| 2006-2010 | 29 (40.8) | 20 (38.5) |
| 2011-2015 | 9 (12.7) | 7 (13.5) |
| 2016-2023 | 4 (5.6) | 1 (1.9) |
| Sample size <sup>a</sup>, median (IQR) | 191 (85-433) | 324 (134-524) |
| Race and ethnicity reported <sup>a</sup> |  |  |
| No race and ethnicity reported | 35 (49.3) | 18 (34.6) |
| Reported Asian/Pacific Islander | 11 (15.5) | 10 (19.2) |
| Reported Black (ethnicity unspecified) | 10 (14.1) | 9 (17.3) |
| Reported Black (non-Hispanic) | 10 (14.1) | 10 (19.2) |
| Reported Hispanic | 13 (18.3) | 12 (23.1) |
| Reported Native American | 2 (2.8) | 1 (1.9) |
| Reported White (ethnicity unspecified) | 25 (35.2) | 23 (44.2) |
| Reported White (non-Hispanic) | 11 (15.5) | 11 (21.2) |
| No. of racial and ethnic group reported <sup>a</sup>, median (IQR) | 1 (0-3) | 1 (0-4) |
| Percentage of Asian/Pacific Islander patients <sup>b</sup>, median (IQR) | 0.9 (0.6-1.6) | 0.9 (0.6-1.6) |
| Percentage of Black (ethnicity unspecified) patients <sup>b</sup>, median (IQR) | 4.5 (3.6-6.6) | 4.6 (4.1-6.6) |
| Percentage of Black (non-Hispanic) patients <sup>b</sup>, median (IQR) | 7.2 (3.7-9.1) | 7.2 (3.7-9.1) |
| Percentage of Hispanic patients <sup>b</sup>, median (IQR) | 5.2 (3.1-6.6) | 5.6 (2.5-6.6) |
| Percentage of Native American patients <sup>b</sup>, median (IQR) | 0.4 (0-0.8) | 0.8 (0.8-0.8) |
| Percentage of White (ethnicity unspecified) patients <sup>b</sup>, median (IQR) | 91.3 (87.3-93.6) | 91.3 (87.3-93.6) |
| Percentage of White (non-Hispanic) patients <sup>b</sup>, median (IQR) | 84.1 (80.1-93.4) | 84.1 (80.1-93.4) |
| Reporting sample characteristics by racial and ethnic groups <sup>a</sup> |  |  |
| No | 71 (100) | 52 (100) |
| Yes | 0 (0) | 0 (0) |
| Reporting detailed trial outcomes (e.g., safety, efficacy) by racial and ethnic groups <sup>a</sup> |  |  |
| No | 71 (100) | 52 (100) |
| Yes | 0 (0) | 0 (0) |
| Analyzing/discussing heterogeneity or differences across racial and ethnic groups <sup>a</sup> |  |  |
| No | 68 (95.8) | 49 (94.2) |
| Yes | 3 (4.2) | 3 (5.8) |
| Discussing the racial and ethnic representation of enrolled trial patients <sup>a</sup> |  |  |
| No | 71 (100) | 52 (100) |
| Yes | 0 (0) | 0 (0) |

Abbreviation: IQR, Interquartile Range.
<sup>a</sup> Among trials with any published data, N=71 for column 2 (any publications), N=52 for column 3 (peer-reviewed publications)
<sup>b</sup> Among trials with reported data on race and ethnicity. The sample size for each reported race and ethnicity is provided above in the table under the variable “Race and ethnicity reported”.

Terminology used to describe racial and ethnic groups varied widely and was often inconsistent with guidelines. Over time, the proportion of trials reporting any race or ethnicity data did not improve, though the number of groups reported slightly increased. (**Figure, A-B**)

**Figure.**
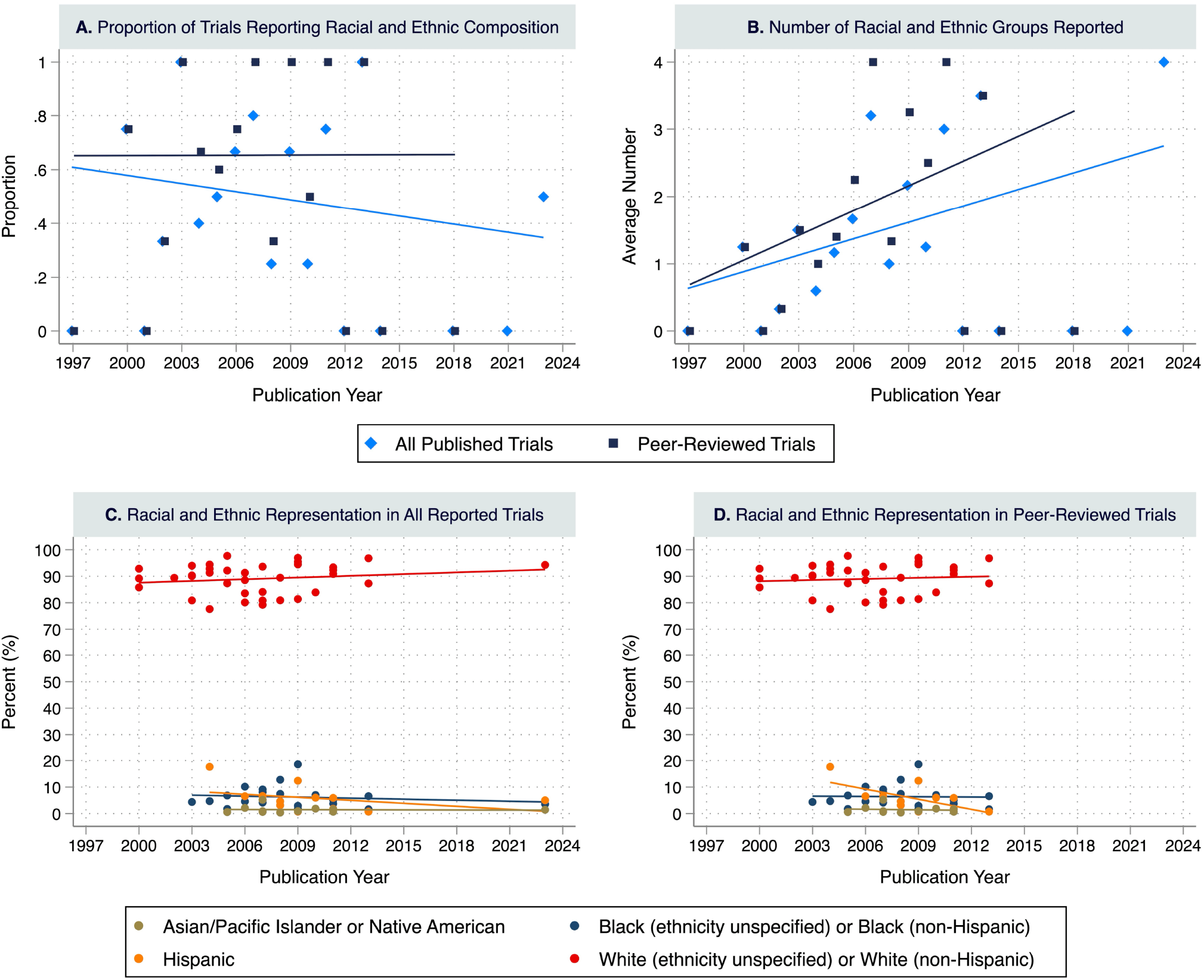
Trends in Racial and Ethnic Reporting and Representation Among US-based Phase III Alzheimer’s Disease Trials, 1997-2023 *Notes*: Panels A and B present the time trend of race and ethnicity reporting over years. Panel A presents the proportion of trials that reported racial and ethnic composition of the study patients. Each dotted points represent the proportion for each year, with light blue representing the proportion among all published trials, and dark blue representing the proportion among trials with peer-reviewed publications. Linear time trends were fitted respectively for all published trials (light blue line) and all peer-reviewed trials (dark blue line). Panel B presents the average number of racial and ethnic groups reported for each year among trials with published data. Estimate for each year were provided as dotted points and linear time trends were fitted respectively for all published trials (light blue color) and peer-reviewed trials (dark blue color). Panels C and D present the racial and ethnic representation among all published trials (Panel C) and peer-reviewed trials (Panel D). Each dotted point represents the percentage of patients from a specific racial and ethnic group in individual clinical trials that reported data on that group. Green dots represent the reported percentages of Asian or Pacific Islander or Native American patients; blue dots represent the reported percentages of Black (ethnicity unspecified) or Black (non-Hispanic) patients; orange dots represent the reported percentages of Hispanic patients; and red dots represent the reported percentages of White (ethnicity unspecified) or White (non-Hispanic) patients. Linear time trends were fitted for each racial and ethnic groups with consistent colors as the dotted points.

When reported, patients from racial and ethnic minoritized groups remained underrepresented: median enrollment was 0.9% (Interquartile Range [IQR]: 0.6–1.6) for Asian or Pacific Islander, 4.5% (IQR: 3.6–6.6) for Black (unspecified ethnicity), 7.2% (IQR: 3.7–9.1) for Black (non-Hispanic), 5.2% (IQR: 3.1–6.6) for Hispanic, and 0.4% (IQR: 0–0.8) for Native American patients (**Table**). No trials acknowledged underrepresentation issue or proposed corrective measures. Minoritized patient enrollment remained consistently low over time, with White patients constituting nearly 90% of trial populations and a slight decline in Hispanic representation (**Figure, C-D**).

## Discussion

This study reveals persistent gaps in the reporting and representation of racial and ethnic minoritized populations in US-based Phase III AD trials. Nearly half of trials failed to report patient race or ethnicity; and when reported, these populations remained consistently underrepresented. Few trials conducted subgroup analyses by race or ethnicity, likely due to low enrollment, limiting the ability to assess treatment safety or efficacy across diverse groups.^2,4,6^

These patterns suggest broader structural and trial-level barriers to equitable inclusion, such as restrictive eligibility criteria, recruitment biases, site selection, and limited community engagement.^1–6^ Addressing these drivers is essential to developing targeted solutions. Future research, particularly using mixed-methods approaches, should explore the underlying causes of low enrollment, including language barriers, cultural factors, mistrust, and trial accessibility.^1–6^

Improving representation will require standardized race and ethnicity reporting, with consistent definitions and mandatory registry fields. Effective recruitment strategies, such as partnering with local advocacy groups and leaders, employing bilingual staff, and tailoring materials for underserved communities, should be adopted more widely to ensure inclusive and generalizable AD research.^1–6^

## Data Availability

All data produced in the present study are available upon reasonable request to the authors.

## Author Contributions

Dr. Chen had full access to all of the data in the study and takes responsibility for the integrity of the data and the accuracy of the data analysis.

*Concept and design*: Lin, Sun, Ross, Chen.

*Acquisition, analysis, or interpretation of data*: All authors.

*Drafting of the manuscript*: Lin, Chen.

*Critical review of the manuscript for important intellectual content*: Lin, Sun, Ross, Chen.

*Statistical analysis*: Lin, Chen.

*Obtained funding*: Ross, Chen.

*Administrative, technical, or material support*: Lin, Chen.

*Supervision*: Lin, Chen.

## Supplementary Online Content

**Supplementary eFigure.**
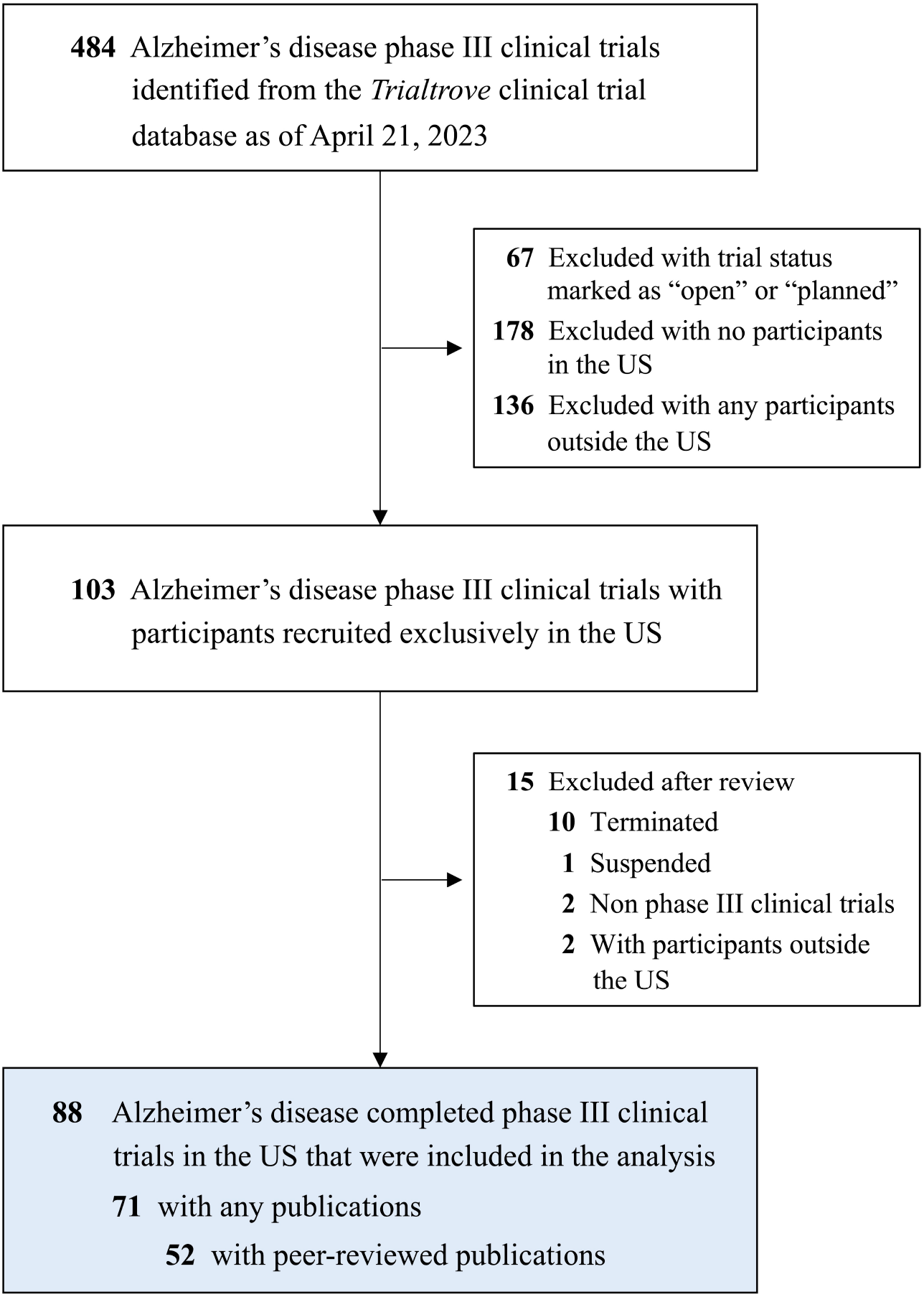
Study Sample Selection Process

